# Nasopharyngeal Panbio COVID-19 antigen performed at point-of-care has a high sensitivity in symptomatic and asymptomatic patients with higher risk for transmission and older age

**DOI:** 10.1101/2020.11.16.20230003

**Authors:** Mar Masiá, Marta Fernández-González, Manuel Sánchez, Mar Carvajal, José Alberto García, Nieves Gonzalo, Victoria Ortiz de la Tabla, Vanesa Agulló, Inmaculada Candela, Jorge Guijarro, José Antonio Gutiérrez, Carlos de Gregorio, Félix Gutiérrez

## Abstract

**Background:** Performance of point-of-care tests in clinical practice remains undetermined. We aimed to evaluate the performance of the nasopharyngeal Panbio COVID-19 antigen Rapid Test Device in real-life conditions in different clinical scenarios.

**Method:** Prospective study conducted in three primary care centers (PCC) and an emergency department. The antigen test was performed at point-of-care in nasopharyngeal and nasal swabs, and in saliva. Positive and negative percent agreement (PPA, NPA) were calculated with the RT-PCR assay as reference standard.

**Results:** Of 913 patients included, 296 (32.3%) were asymptomatic and 690 (75.6%) came from the PCC. Nasopharyngeal swabs were collected from 913, nasal swabs from 659, and saliva from 611 patients. RT-PCR was positive in 196 (21.5%) nasopharyngeal samples (NPS). Overall PPA (95% CI) in NPS was 60.5% (53.3-67.4), and it was lower in nasal swabs (44.7%) and saliva (23.1%). Test performance in NPS was largely dependent on the cycle threshold (Ct) in RT-PCR, with PPA>90% for Ct≤25 and ≥80% for Ct<30. In symptomatic patients, the PPA was 95% for Ct≤25; ≥85% for Ct<30, and 89% for the symptom triad of fever, cough and malaise. Performance was also dependent on age, with PPA of 100% in symptomatic patients >50 years with Ct<25. In asymptomatic patients, the PPA was 86% for Ct<25. In all cases, NPA was 100%.

**Conclusion:** The nasopharyngeal Panbio COVID-19 antigen test performed at point-of-care is highly sensitive in symptomatic patients, particularly with Ct<30 and older age. The test was useful to identify asymptomatic patients with lower Ct values and therefore with contagious risk.

**Key points:** The nasopharyngeal Panbio-COVID-19 antigen test performed in real-life conditions at point-of-care is highly sensitive in symptomatic patients, particularly with Ct<30 and older age. The test is useful to identify asymptomatic patients with lower Ct values and therefore with contagious risk.

---

The rapid spread of the SARS-CoV-2 pandemic worldwide requires the urgent adoption of effective preventive measures. Early diagnosis and rapid isolation of infected people are central to contain disease transmission. While real-time reverse transcription polymerase chain reaction (RT-PCR) is currently the reference assay for the diagnosis of SARS-CoV-2 infection, novel rapid antigen tests have emerged with several potential key advantages over molecular methods [1]. In contrast to the RT-PCR, the antigen test is relatively inexpensive, simple to perform and easy to interpret, does not require infrastructure, and enables obtaining point-of-care results within a few minutes. As a result, it allows immediate decisions about isolation and therapeutic interventions on infected individuals. Moreover, antigen tests are capable of identifying infected people early after infection, when viral loads are high and the likelihood of transmission is highest [2]. Despite the lower sensitivity when compared to the molecular assays, the possibility of repetitive testing with a low cost procedure and the real-time detection of the most infective patients, make the antigen a potentially high valuable test in terms of surveillance, to track and prevent the spread of the infection [3].

Information on the performance of the point-of-care SARS-CoV-2 antigen tests is limited. The sensitivity of the first generation antigens is overall low [4], and most studies have been conducted in laboratory specimens, involved a relatively low number of samples, and the minority with available clinical data included primarily symptomatic patients [5-8]. To assess the real performance of a point-of-care test, it should be used in real-life conditions, including consecutive patients, and obtaining results on site. One of the uncertainties about the antigen is the accuracy of the test in asymptomatic patients, and how it performs in additional clinical settings, like childhood or old age, among others. Another relevant question is whether a more convenient sample would be a suitable alternative for diagnosis. Antigen tests are currently authorized to be performed on nasopharyngeal or nasal swabs, which need to be collected by healthcare professionals. Since saliva can be self-collected, antigen assessment in this sample would facilitate large-scale testing.

Panbio COVID-19 antigen Rapid Test Device (RTD) (Abbott Rapid Diagnostic Jena GmbH, Jena, Germany) has been recently marketed for the qualitative detection of SARS-CoV-2 antigen in human nasopharyngeal swab specimens, with high sensitivity and specificity. We evaluated the performance of this point-of-care test in real-life conditions, in three Primary Care Centers (PCC) and an Emergency Department (ED). We assessed the accuracy of the test in symptomatic and asymptomatic patients, in different clinical scenarios, and in nasopharyngeal, nasal and saliva samples.

## METHODS

### Study design, setting and data collection

A prospective study was conducted from 15^th^ September to 29^th^ October 2020 in three PCC and an ED. Consecutive patients, either with COVID-19 signs/symptoms or asymptomatic contacts attending the PCC, and a majority of symptomatic patients presenting to the ED were included in the study. Demographic and clinical data from primary care patients were collected using a structured questionnaire. The questionnaire included information about six specific symptoms and their temporality and the number of days since the initiation of symptoms. Clinical data from patients who attended the ED were obtained from the electronic health records. Informed consent was obtained from all patients, and the study was approved by the Hospital General Universitario de Elche COVID-19 Institutional Advisory Board.

### Specimen collection

At the PCC, patients were asked to fill the questionnaire about symptoms and to collect saliva into a 100 ml sterile empty container. Then, a nasal swab and two consecutive nasopharyngeal swabs (one swab for each nostril) were obtained by a qualified nurse according to the recommended standard procedure. At the ED, two consecutive nasopharyngeal swabs, also with a different swab for each nostril, were obtained by a clinician.

### Microbiological procedures

#### Antigen detection

Nasal swabs, one of the two nasopharyngeal swabs and the saliva samples obtained at the PCC were tested on-site in the following minutes after collection for antigen detection. One of the nasopharyngeal swabs obtained at the ED was also analyzed on-site for antigen detection immediately after collection. The antigenic assessment in all the samples was performed using the Panbio COVID-19 Ag RTD, an immunochromatographic test with a membrane strip pre-coated with antibodies to the SARS CoV-2 nucleocapsid. The kit was used according to the manufacturer’s instructions. Briefly, nasal and nasopharyngeal swabs were immersed in two extraction tubes containing 300 µl of buffer from the kit. A third swab was soaked in the saliva sample and then immersed in a third 300 µL-tube. The three tubes were ready to be applied to the corresponding antigen device.

#### SARS-CoV-2 RNA detection

The second nasopharyngeal swab was preserved in a 3 mL transport tube containing guanidine salt solution (Mole Bioscience, SUNGO Europe B.V., Amsterdam, Netherlands). After collection of all samples, nasopharyngeal specimens were transported daily by the same healthcare workers who collected the samples at the PCC to the clinical microbiology laboratory for immediate molecular analysis by RT-PCR. Nasopharyngeal samples (NPS) from the ED were also sent to the same microbiology laboratory. Nucleic acid extraction was performed using 300 µL of nasopharyngeal specimen on Chemagic™ 360 Nucleic Acid Purification Instrument (PerkinElmer España SL, Madrid, Spain). Then, 10 uL of eluate was used for real-time RT-PCR assay targeting the E-gene (LightMix® Modular SARS-CoV (COVID19) E gene, TIB MOLBIOL, Berlin, Germany, distributed by Roche). Testing was performed according to the manufacturer’s guidelines on Cobas z 480 Analyzer (Roche, Basilea, Suiza).

### Statistical analyses

The percent agreement (positive, negative and overall) (PPA, NPA and OPA) for Panbio antigen test in the nasopharyngeal, nasal and saliva samples compared to the reference standard RT-RCR test in nasopharyngeal swab was calculated. Performance agreement was evaluated using Cohen’s kappa coefficient. Performance was also evaluated in NPS stratifying by age, sex, the number of cycles of amplification in RT-PCR (cycle threshold, Ct), and duration of symptoms. Multivariate logistic regression was performed to assess predictors of the sensitivity of the antigen test in symptomatic patients. The estimated sample size for a sensitivity of at least 91.4% (according to the manufacturer), a precision of 2.5% and a statistical power of 80%, was 762 patients. For a specificity of at least 97%, sample size required was 377 patients.

## RESULTS

During the study period, 913 patients were included; all of them had a nasopharyngeal swab for RT-PCR, 904 (99%) a second nasopharyngeal swab for antigen test, 659 (72%) a nasal swab and 611 (67%) a saliva sample collected. A total of 690 (75.6) NP samples were collected from the PCC and 223 (24.4) from the ED.

Clinical characteristics of the patients are shown in Table 1. Median (Q1-Q3) age was 40.6 (23.0-55.6) years, and 423 (46.3%) were men. The most common comorbidities were dyslipidemia in 80 (22.2%) patients, hypertension in 124 (17.0%), and diabetes in 60 (8.2%). There were 617 (67.6%) symptomatic patients and 296 (32.4%) were asymptomatic. Median (Q1-Q3) number of days from symptom onset was 3 (2-5) days, and the most frequent symptoms were cough (50.1%), followed by fever (46.8%), sore throat (31.9%) and nasal congestion (31.3%). Median (Q1-Q3) Ct was 24 (16-30); 22 (16-29) in symptomatic and 28 (21-32) in asymptomatic patients (p=0.012); and 21 (15-27), in patients ≥50 years and 26 (18-31) in <50 years (p=0.02).

**Table 1.** Clinical characteristics of the patients

| Variable | All patients<br>N=913 | Symptomatic<br>N=617 | Asymptomatic<br>N=296 | P |
| --- | --- | --- | --- | --- |
| Age (years) median (Q1-Q3) | 40.6 (23-55.6) | 41 (24-56.3) | 39.9 (20.4-52.5) | 0.086 |
| Sex (men) | 423 (46.3) | 289 (46.8) | 134 (45.3) | 0.671 |
| Dyslipidemia | 80 (22.2) | 63 (24.3) | 17 (16.7) | 0.124 |
| Hypertension | 124 (17.0) | 96 (19.0) | 28 (12.4) | 0.033 |
| Diabetes | 60 (8.2) | 49 (9.7) | 11 (4.9) | 0.029 |
| Cardiomyopathy | 55 (7.6) | 43 (8.5) | 12 (5.3) | 0.171 |
| Obesity | 42 (5.8) | 25 (5.1) | 17 (7.6) | 0.228 |
| COPD | 25 (3.4) | 20 (4.0) | 5 (2.2) | 0.277 |
| Active cancer | 17 (2.3) | 14 (2.8) | 3 (1.3) | 0.295 |
| Primary care center | 690 (75.6) | 416 (67.4) | 274 (92.6) | <0.001 |
| Emergency department | 223 (24.4) | 201 (32.6) | 22 (7.4) | <0.001 |
| Positive SARS-Cov-2 RNA * | 196 (21.5) | 156 (25.3) | 40 (13.5) | <0.001 |
| Ct, median (Q1-Q3) | 24 (16-30) | 22 (16-29) | 28 (21-32) | 0.012 |
| Ct 0-20 | 78 (40.0) | 68 (43.9) | 10 (25.0) | 0.10 |
| Ct 21-25 | 29 (14.9) | 24 (15.5) | 5 (12.5) |  |
| Ct 26-30 | 40 (20.5) | 28 (18.1) | 12 (30.0) |  |
| Ct 31-35 | 40 (20.5) | 30 (19.4) | 10 (25.0) |  |
| Ct >35 | 8 (4.1) | 5 (3.2) | 3 (7.5) |  |
| Positive antigen (any NP-nasal-saliva) result | 120 (13.1) | 106 (17.2) | 14 (4.7) | <0.001 |
| Positive NP antigen result | 118 (12.9) | 105 (17.2) | 13 (4.5) | <0.001 |
| No. of days with symptoms |  | 3 (2-5) |  |  |
| Cough |  | 309 (50.1) |  |  |
| Fever |  | 289 (46.8) |  |  |
| Sore throat |  | 196 (31.9) |  |  |
| Nasal congestion |  | 193 (31.3) |  |  |
| Dyspnea |  | 115 (18.6) |  |  |
| Anosmia/ageusia |  | 53(8.8)/44(7.1) |  |  |
| Others: malaise/headache |  | 104(17)/140(23) |  |  |

Categorical variables are represented by number and (%). \*Performed on nasopharyngeal samples. COPD, chronic obstructive pulmonary disease; Ct, cycle threshold of RT-PCR; NP, nasopharyngeal;

### Performance of the antigen test by type of sample, site of care, and cycle threshold

There were 196 (21.5%) samples positive for SARS-CoV-2 by RT-PCR and 120 (13.1%) positive antigen results. Performance of the test by type of sample is shown in Figure 1 and Table 2. In NPS, the overall PPA and NPA (95% CI) of the antigen test were 60.5% (53.3-67.4) and 100% (99.3-100), respectively. In the saliva, the PPA (95% CI) was 23.1% (16.2-31.9), and in nasal samples 44.7% (36.1-53.6).

**Table 2.** Performance of the Panbio COVID-19 antigen Rapid Test Device

| **Overall** | TP | FP | TN | FN | PPA (95% CI) | NPA (95% CI) | OPA (95% CI) | Kappa (95% CI) |
| --- | --- | --- | --- | --- | --- | --- | --- | --- |
| NP sample | 118 | 0 | 709 | 77 | 60.5% (53.3-67.4) | 100% (99.3-100) | 91.5% (89.4-93.2) | 0.71 (0.65-0.77) |
| Nasal sample | 59 | 0 | 527 | 73 | 44.7% (36.1-53.6) | 100% (99.1-100) | 88.9% (86.2-91.2) | 0.56 (0.48-0.65) |
| Saliva sample | 28 | 0 | 490 | 93 | 23.1% (16.2-31.9) | 100% (99-100) | 84.8% (81.6-87.59) | 0.33 (0.23-0.42) |
| **Site of care** |  |  |  |  |  |  |  |  |
| Primary care | 78 | 0 | 544 | 59 | 56.9% (48.2-65.3) | 100% (99.1-100) | 91.3% (88.9-93.3) | 0.68 (0.60-0.75) |
| Emergency department | 40 | 0 | 165 | 18 | 69% (55.3-80.1) | 100% (97.2-100) | 91.9% (87.3-95) | 0.77 (0.67-0.87) |
| **Sex** |  |  |  |  |  |  |  |  |
| Men | 64 | 0 | 317 | 37 | 63.4% (53.1-72.6) | 100% (98.5-100) | 91.1% (87.9-93.6) | 0.72 (0.64-0.81) |
| Women | 54 | 0 | 392 | 40 | 57.4% (46.8-67.5) | 100% (98.8-100) | 91.8% (88.9-94) | 0.69 (0.60-0.77) |
| **Age** |  |  |  |  |  |  |  |  |
| ≤14 y | 10 | 0 | 107 | 8 | 55.6% (31.3-77.6) | 100% (95.7-100) | 93.6% (87.4-97) | 0.68 (0.48-0.88) |
| 15-30 y | 16 | 0 | 145 | 26 | 38.1% (24-54.3) | 100% (96.8-100) | 86.1% (80.1-90.6) | 0.49 (0.33-0.65) |
| 31-50 y | 42 | 0 | 244 | 27 | 60.9% (48.4-72.2) | 100% (98.1-100) | 91.4 (87.6-94.1) | 0.71 (0.61-0.81) |
| 51-65 y | 29 | 0 | 111 | 8 | 78.4% (61.3-89.6) | 100% (95.8-100) | 94.6% (89.3-97.5) | 0.85 (0.74-0.95) |
| >65 y | 21 | 0 | 102 | 8 | 72.4% (52.5-86.6) | 100% (95.5-100) | 93.9% (87.9-97.1) | 0.80 (0.67-0.93) |
| **Symptomatic** |  |  |  |  |  |  |  |  |
| Overall | 105 | 0 | 456 | 51 | 67.3% (59.3-74.5) | 100% (99-100) | 91.7% (89.1-93.7) | 0.75 (0.69-0.82) |
| ≤ 3 DSO | 49 | 0 | 273 | 13 | 79% (66.5-87.9) | 100% (98.3-100) | 96.1% (93.3-97.8) | 0.86 (0.79-0.93) |
| ≤ 4 DSO | 65 | 0 | 334 | 18 | 78.3% (67.6-86.3) | 100% (98.6-100) | 95.7% (93.1-97.3) | 0.85 (0.79-0.92) |
| ≤ 5 DSO | 76 | 0 | 364 | 22 | 77.6% (67.8-85.1) | 100% (98.7-100) | 95.2% (92.8-96.9) | 0.85 (0.78-0.91) |
| ≤ 6 DSO | 81 | 0 | 382 | 23 | 77.9% (68.5-85.2) | 100% (98.8-100) | 95.3% (92.9-96.9) | 0.85 (0.79-0.91) |
| ≥7 DSO | 24 | 0 | 74 | 28 | 46.2% (32.5-60.4) | 100 (93.9-100) | 77.8% (69.3-84.5) | 0.50 (0.36-0.64) |
| Ct ≤ 25 | 87 | 0 | 0 | 5 | 95% (87-98) |  |  |  |
| Ct ≤ 30 | 102 | 0 | 0 | 18 | 85% (77-91) |  |  |  |
| Ct ≤ 35 | 105 | 0 | 0 | 45 | 70% (62-77) |  |  |  |
| **Asymptomatic** |  |  |  |  |  |  |  |  |
| Overall | 13 | 0 | 253 | 26 | 33.3% (19.6-50.3) | 100% (98.1-100) | 91.1% (87.1-94) | 0.46 (0.30-0.63) |
| Ct ≤ 25 | 12 | 0 | 0 | 2 | 86% (56-97) |  |  |  |
| Ct ≤ 30 | 13 | 0 | 0 | 13 | 50% (32-68) |  |  |  |
| Ct ≤ 35 | 13 | 0 | 0 | 23 | 36% (21-54) |  |  |  |

Unless specified, all analyses have been performed in nasopharyngeal samples. TP, true positive; FP, false positive; TN, true negative; FN, false negative; PPA, positive percent agreement; NPA, negative percent agreement; OPA, overall predictive agreement; NP, nasopharyngeal; y, year; DSO, days from symptom onset; Ct, cycle threshold of RT-PCR

**Figure 1.**
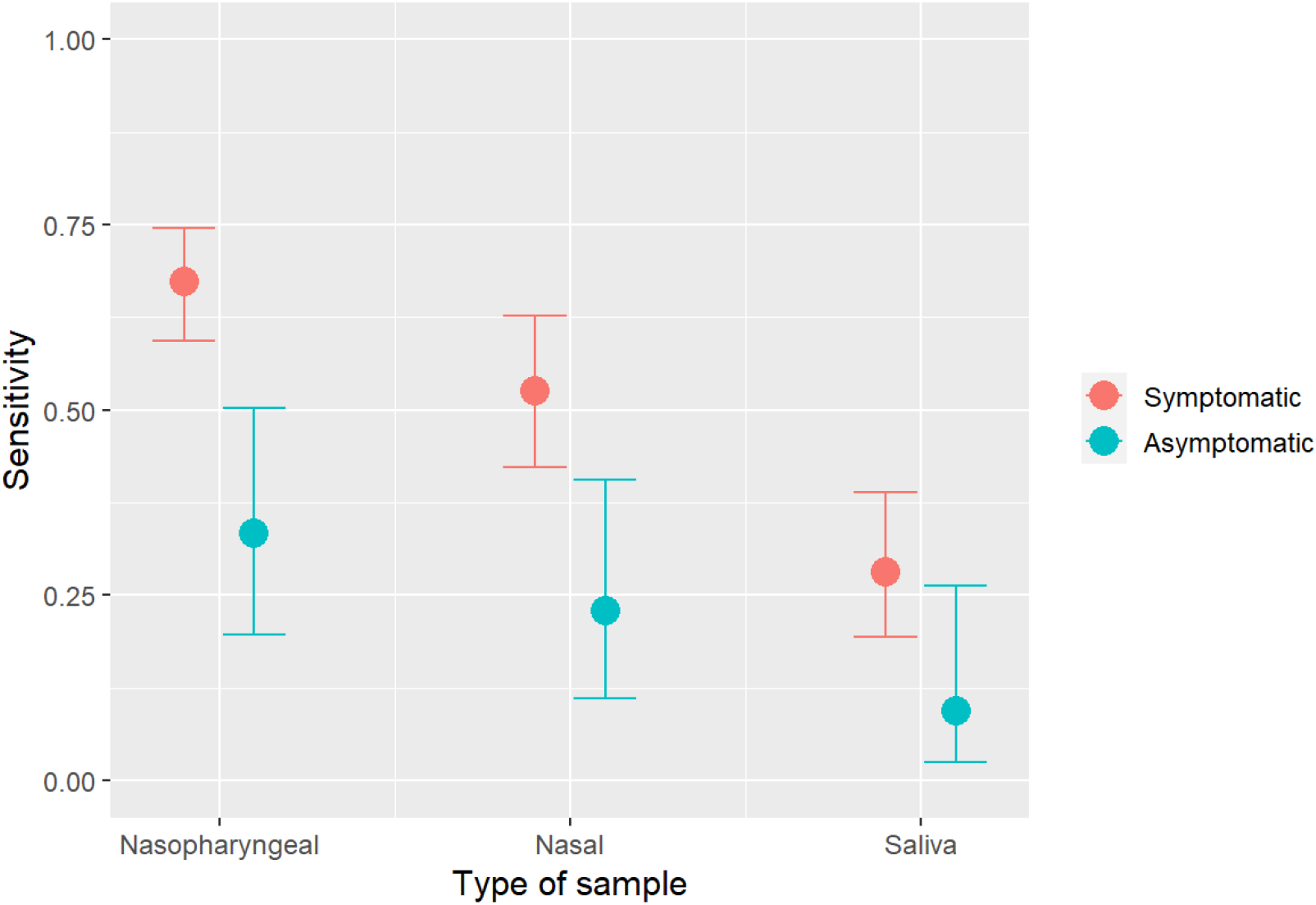
Performance of nasopharyngeal Panbio COVID-19 antigen Rapid Test Device by type of sample

By site of care, the PPA (95% CI) in NPS at the ED was 69.0% (55.3-80.1). At the PCC, the PPA was 56.9% (48.2-65.3).

The performance of the test by Ct in NPS is shown in Figure 2A. In the analysis including all patients, a gradual decline in sensitivity was observed with increasing Ct values, with a more prominent decrease from Ct>28. The PPA was >90% for Ct ≤25, and ≥80% for Ct <30.

**Figure 2.**
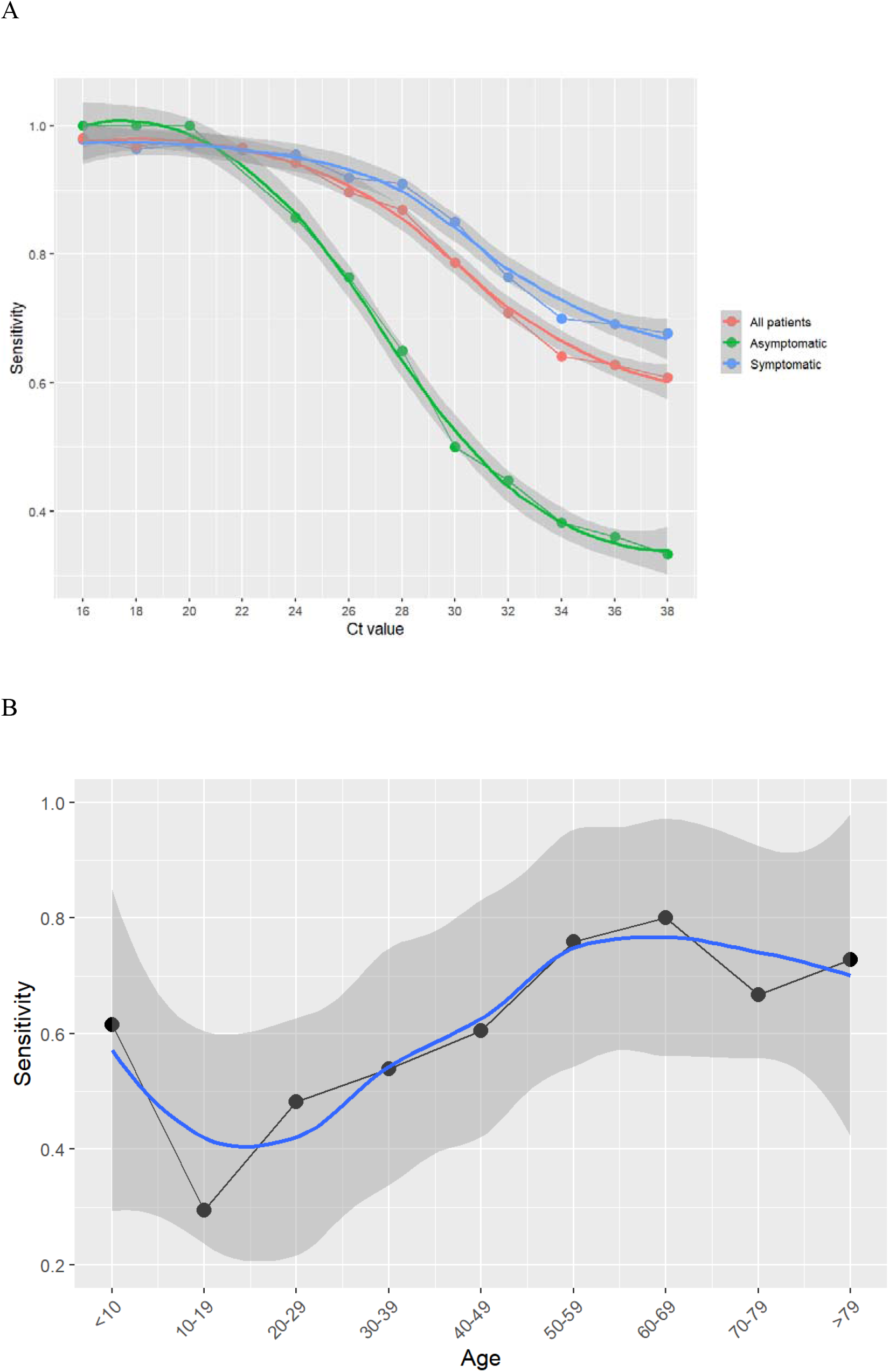
Performance of nasopharyngeal Panbio COVID-19 antigen Rapid Test Device in different scenarios. A: Performance according to Ct values. B: Performance according to age.

### Performance of the antigen test in NPS by age and sex

Table 2 and Figure 2B show the performance of the antigen test according to age group. There PPA increased with increasing age, with lower sensitivity among children and young adults (PPA [95% CI] 38.1% [24.0-54.3] for 15-30 years,) and higher sensitivity in older patients (PPA [95% CI] 72.4% [52.5-86.6] for ≥65 years).

No remarkable differences were found in the antigen test performance by sex. The PPA in men was 63.4% (53.1-72.6), and in women, 57.4% (46.8-67.5) (Table 2).

### Performance of the antigen test in NPS in symptomatic patients

A total of 617 (67.6%) patients presented with clinical symptoms. Median (Q1-Q3) age was 41.0 (24.0-56.3) years and 289 (46.8%) were men (Table 1). In 156 (25.3%) patients the RT-PCR was positive in the nasopharyngeal sample and in 105 (17.2%) the antigen test was positive. The PPA (95% CI) in the NPS samples was 67.3% (59.3-74.5). The PPA by type of sample in symptomatic patients is shown in Figure 1.

The performance of the antigen test in NPS by Ct is shown in Figure 2A and Table 2. The sensitivity of the test decreased more slowly with increasing Ct in symptomatic patients than in the overall sample, although a faster decrease was again observed from Ct values >28. The PPA was ≥95% (95% CI 87-98) for Ct ≤25, and ≥85% (77-91) for Ct <30.

By age, the antigen test performance increased with increasing age, with a PPA (95% CI) of 79.3% (66.3-88.4) in patients >50 years.

Table 2 shows the performance of the antigen test in NPS by number of days with symptoms. The PPA was near 80% for a period <7 days from symptom onset, and fell from then on.

By symptoms, the highest sensitivity was observed for malaise and ageusia, with a PPA of 75% each, followed by sore throat with PPA 73%, and cough, nasal congestion and dyspnea, the three with PPA of 69%. The PPA for the triad of cough, fever and malaise was 89%.

Figure 3 shows the performance of the antigen test according to the presence of symptoms, age, Ct and days after symptoms onset. The highest PPA of the test was observed for Ct<25, for which the PPA was >90% for concomitant age >15 years; 100% for >50 years; and 95% for <7 days duration of symptoms.

**Figure 3.**
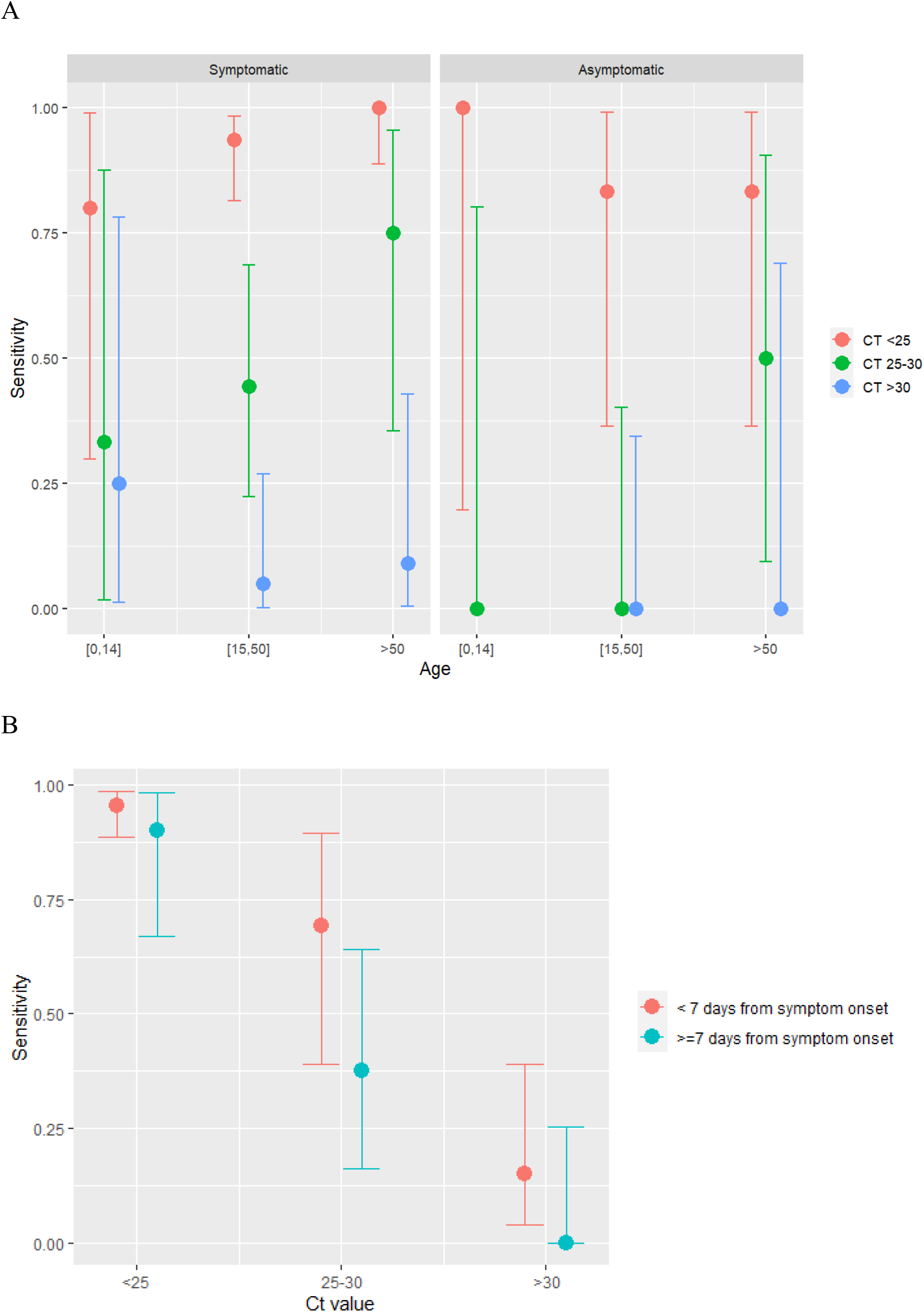
Performance of nasopharyngeal Panbio COVID-19 antigen Rapid Test Device after stratification for different factors. A: Performance stratified by the presence of symptoms, age, and Ct values. B: Performance stratified by symptom duration and age.

A multivariate logistic regression was run to explore the independent factors associated with antigen test performance among symptomatic patients, including age, sex, Ct values and duration of symptoms categorized into < or ≥7 days. The model showed that the PPA was independently associated with age, with an OR (95% CI) of 1.20 (1.03 - 1.40) for each 5 year-older period, duration of symptoms with an OR of 3.99 (1.22-13.06) for <7 days, and inversely associated with the Ct, with an OR (95% CI) of 0.66 (0.57-0.76).

### Performance of the antigen test in NPS in asymptomatic patients

A total of 296 patients were asymptomatic, with median (Q1-Q3) age of 39.9 (20.4-52.5) years and 134 (45.3%) were men. The PPA by type of sample in asymptomatic patients is shown in Figure 1. A total of 39 (13.2%) patients had a positive RT-PCR in the nasopharyngeal sample and 13 (4.4%) a positive NP antigen test. The PPA in the nasopharyngeal sample was 33.3% (95% CI, 19.6-50.3), and Cohen’s kappa coefficient 0.46 (95% CI, 0.30-0.63).

Figure 2A, Fig. 3A and Table 2 show the performance of the antigen test by Ct and age in asymptomatic patients. Again a decrease in sensitivity was observed with increasing Ct, but it was much more pronounced than in symptomatic patients, mainly from Ct>20 (Fig. 2A). However, for low Ct, the sensitivity was high for all age groups, with an overall PPA (95% CI) of 86% (56-97) for Ct ≤25.

## DISCUSSION

We evaluated a recent generation point-of-care antigen test for SARS-CoV-2 in real-life conditions in a large population of consecutive patients and on-site, where the test was conceived to be performed. Our data show that the sensitivity of the antigen test is largely dependent on the Ct values, age, and the presence and duration of symptoms. The sensitivity of the antigen test was highest in symptomatic patients older than 50 years and with Ct values associated with an increased risk of infectivity, reaching 100% in this scenario. Although the performance of the test was overall lower in asymptomatic patients, again the antigen identified with a sensitivity higher than 85% those with lower Ct, and therefore with higher contagious risk. In all cases, the specificity of the antigen test was near 100%. Finally, although the saliva would facilitate mass testing for surveillance, the low sensitivity of the antigen in this specimen does not support its use as an alternative sample.

In contrast to SARS-CoV-1, SARS-CoV-2 infection is associated with high levels of viral shedding at the initial stages of the infection in the upper respiratory tract, which facilitates detecting the virus during the most infectious period. The availability of a rapid point-of-care test for the diagnosis allows adopting immediate and real-time decisions, which is a clear advantage over the RT-PCR in controlling the spread of the infection. Although the antigen test showed an overall lower sensitivity than the RT-PCR in our study, and that reported by the manufacturer, the test was highly accurate in symptomatic patients exhibiting lower Ct values, with a sensitivity greater than 95% for Ct of 25 or lower, and at least 85% for Ct of less than 30. High SARS-CoV-2 viral load has been associated with severity of disease and mortality [9,10], and several studies support a correlation of Ct values with infectivity, as defined by growth in cell culture [11-13]. Although breakpoints fluctuate among different studies, a diagnostic Ct value of RT-PCR equal or greater than 32 was associated with no isolation of SARS-CoV-2 using cell-based cultures, neither with active viral replication [11]. Other studies report no SARS-CoV-2 recovery in cell culture with Ct values higher than 29.5 [12], or even higher than 24, with a decrease by 32% of positive cultures for each unit of increase in Ct [13]. Although the correlation between viral growth and infectivity needs to be confirmed, our data suggest that the point-of-care antigen test is useful to detect most SARS-CoV-2-infected symptomatic patients, and to identify those with significant transmission risk. Another factor influencing the performance of the antigen test was the duration of symptoms. As specified by the manufacturer and also reported [6], we found a higher sensitivity of the test within a period of less than 7 days from the initiation of symptoms.

Since most SARS-CoV-2 infections are asymptomatic, the performance of the antigen test in this scenario needs to be established, and this information is key for strategies aimed at preventing the spread of the infection at a community level. Our study shows that the sensitivity was poorer when compared to that of patients with symptoms. The same as with symptomatic participants, the sensitivity was highly dependent on the Ct, and we found a PPA higher than 85% for Ct ≤25. Asymptomatic SARS-CoV-2 transmission has been reported [14,15], but the secondary attack rate from either asymptomatic or presymptomatic patients was found in a meta-analysis to be lower compared with that of patients with symptoms [16], and to be as low as 0.3-0.6% [17]. In presymptomatic patients, SARS-CoV-2 growth in viral culture was rarely observed with Ct above 25 [14]. Although the performance of the test was inferior in asymptomatic individuals, the increase in sensitivity observed with lower Ct coupled with the lower transmission risk described within this group, could make the antigen a potentially helpful tool to identify those with infective risk among asymptomatic patients. As with other studies [18], because we did not follow up patients, we cannot distinguish the proportion of asymptomatic SARS-CoV-2 infected individuals who remained asymptomatic throughout, and those who were presymptomatic and developed symptoms later in the course of the infection. The latter patients may have the chance to be detected by the test in ulterior examinations, thereby increasing the sensitivity of an assay that allows repeated testing because of its inexpensiveness and simplicity.

In addition to the viral load, the sensitivity of the antigen test was highly dependent on age. Younger children showed the poorest antigen test performance, and there was a gradual increase in the sensitivity of the test with age, with highest values in older patients. Several factors might contribute to explain this finding. Children showed less cooperation or even resistance during the collection of the nasopharyngeal sample. Other factors like temporality of symptoms or the higher Ct values among younger patients might also have played a role. However, our study showed that age was associated with the antigen test performance independently of the Ct and duration of symptoms, a finding that merits further investigation.

We explored the performance of the antigen test in alternative locations to the nasopharynx recommended by the manufacturer, which could be more useful for surveillance, like the nose or saliva. Because saliva can be self-collected, this sample would be the most advantageous if mass testing was considered with the point-of-care antigen test. Saliva has additionally shown to be a suitable alternative sample to nasopharyngeal swab for SARS-CoV-2 detection by RT-PCR [19]. Unfortunately, our study shows that the sensitivity of the point-of-care antigen test is low in the saliva. The same occurred with the nasal swabs, where the test performance was neither satisfactory.

Limitations of the study include the lack of statistical power for the analysis of the test performance in specific subgroups, and the incomplete information about number of days since the risk contact in asymptomatic patients. Strengths are the real-life conditions in which the antigen test has been used to assess its true performance, the inclusion of consecutive unselected patients which allowed analyzing how it performs in diverse clinical scenarios, and the on-site execution of the test.

In conclusion, the sensitivity of the nasopharyngeal Panbio COVID-19 antigen RDT is closely related to the Ct values, age and the presence and duration of symptoms. The test performance is optimal in symptomatic patients older than 50 years with viral loads linked with infectivity, and in asymptomatic patients the test may be useful to identify those with a higher risk for transmission. The saliva is not a suitable alternative sample for antigen detection.

## Data Availability

The datasets generated and/or analyzed during the current study are available from the corresponding author on reasonable request.

## Acknowledgements

*Members of the COVID19-Elx-Rapid Diagnostic Tests Study Group*

Félix Gutiérrez, Mar Masiá, Sergio Padilla, Guillermo Telenti, Lucia Guillen, Cristina Bas, María Andreo, Fernando Lidón, Vladimir Ospino, José López, Marta Fernández, Vanesa Agulló, Gabriel Estañ, Javier García, Cristina Martínez, Leticia Alonso, Joan Sanchís, Ángela Botella, Paula Mascarell, María Selene Falcón, Sandra Ruiz, José Carlos Asenjo, Carolina Ding, Mar Carvajal, Inmaculada Candela, Jorge Guijarro, Cristina la Moneda, Cristina Jara, Raquel Mora, Juan Manuel Quinto, Sergio Ros, Daniel Canal, Pascual Pérez, Carolina Garrido, Manuel Sánchez, Jaime Sastre, Carlos de Gregorio, Francisco Carrasco, Juan Navarro, Andrés Navarro, Nieves Gonzalo, Clara Pérez, Adoración Alcalá, José Luis Rincón, Montserrat Ruiz, Juan Antonio Gutiérrez.

## REFERENCES

1. La Marca A, Capuzzo M, Paglia T, Roli L, Trenti T, Nelson SM. Testing for SARS-CoV-2 (COVID-19): a systematic review and clinical guide to molecular and serological in-vitro diagnostic assays. Reprod Biomed Online 2020; 41:483–99.

2. He X, Lau EHY, Wu P, et al. Temporal dynamics in viral shedding and transmissibility of COVID-19. Nat Med 2020; 26: 672–5.

3. Mina MJ, Parker R, Larremore DB. Rethinking Covid-19 Test Sensitivity - A Strategy for Containment. N Engl J Med 2020 [Epub ahead of print].

4. Dinnes J, Deeks JJ, Adriano A, et al. Cochrane COVID-19. Rapid, point-of-care antigen and molecular-based tests for diagnosis of SARS-CoV-2 infection. Cochrane Database Syst Rev 2020; 8:CD013705.

5. Young S, Taylor SN, Cammarata CL, et al. Clinical evaluation of BD Veritor SARS-CoV-2 point-of-care test performance compared to PCR-based testing and versus the Sofia 2 SARS Antigen point-of-care test. J Clin Microbiol 2020 [Epub ahead of print].

6. Linares M, Pérez-Tanoira R, Carrero A, et al. Panbio antigen rapid test is reliable to diagnose SARS-CoV-2 infection in the first 7 days after the onset of symptoms. J Clin Virol 2020; 133:104659.

7. Blairon L, Wilmet A, Beukinga I, Tré-Hardy M. Implementation of rapid SARS-CoV-2 antigenic testing in a laboratory without access to molecular methods: Experiences of a general hospital. J Clin Virol 2020; 129:104472.

8. Lambert-Niclot S, Cuffel A, Le Pape S, et al. Evaluation of a Rapid Diagnostic Assay for Detection of SARS-CoV-2 Antigen in Nasopharyngeal Swabs. J Clin Microbiol 2020; 58:e00977–20.

9. Zheng S, Fan J, Yu F, et al. Viral load dynamics and disease severity in patients infected with SARS-CoV-2 in Zhejiang province, China, January-March 2020: retrospective cohort study. BMJ 2020; 369:m1443.

10. Pujadas E, Chaudhry F, McBride R, et al. SARS-CoV-2 viral load predicts COVID-19 mortality. Lancet Respir Med 2020; 8:e70.

11. Basile K, McPhie K, Carter I, et al. Cell-based culture of SARS-CoV-2 informs infectivity and safe de-isolation assessments during COVID-19. Clin Infect Dis 2020 [Epub ahead of print].

12. Gniazdowski V, Morris CP, Wohl S, et al. Repeat COVID-19 Molecular Testing: Correlation of SARS-CoV-2 Culture with Molecular Assays and Cycle Thresholds. Clin Infect Dis 2020 [Epub ahead of print].

13. Bullard J, Dust K, Funk D, et al. Predicting infectious SARS-CoV-2 from diagnostic samples. Clin Infect Dis 2020 [Epub ahead of print].

14. Arons MM, Hatfield KM, Reddy SC, et al. Presymptomatic SARS-CoV-2 Infections and Transmission in a Skilled Nursing Facility. N Engl J Med 2020; 382:2081–90.

15. Chau NVV, Thanh Lam V, Thanh Dung N, et al. The natural history and transmission potential of asymptomatic SARS-CoV-2 infection. Clin Infect Dis 2020 [Epub ahead of print].

16. Buitrago-Garcia D, Egli-Gany D, Counotte MJ, et al. Occurrence and transmission potential of asymptomatic and presymptomatic SARS-CoV-2 infections: A living systematic review and meta-analysis. PLoS Med 2020; 17:e1003346.

17. Luo L, Liu D, Liao X, et al. Contact Settings and Risk for Transmission in 3410 Close Contacts of Patients With COVID-19 in Guangzhou, China: A Prospective Cohort Study. Ann Intern Med 2020 [Epub ahead of print].

18. Oran DP, Topol EJ. Prevalence of Asymptomatic Sars-Cov-2 Infection: A Narrative Review. Ann Intern Med 2020 [Epub ahead of print].

19. Azzi L, Carcano G., Gianfagna F., Grossi P., Gasperina D.D., Genoni A. Saliva is a reliable tool to detect SARS-CoV-2. J Infect 2020; 81:e45–e50.

